# Types of occupational accidents and their predictors at construction sites in Lusaka city

**DOI:** 10.1101/2022.05.28.22275096

**Authors:** P. Simutenda, M. Zambwe, R. Mutemwa

## Abstract

**Objective:** To determine the types of occupational accidents and their predictors at construction sites in Lusaka city.

**Methods:** A qualitative research design which explored the views, experiences, and beliefs of construction workers. A sample size of seven participants who were previously involved in at least one construction accident was drawn purposively. Data was collected by interviewing participants using a semi-structured interview schedule and a voice recorder. The data from the voice recorder was then transcribed in verbatim format. Thematic evaluation was used for data analysis.

**Findings:** Two themes were identified:- types of occupational accidents and predictors of accidents. The types of construction accidents included crane or hoist accidents, slips, and falls from heights, gas leaks, fires and explosions, forklift, trench, electrocutions, machinery, moving or failing object, caught-between and exposure to dangerous chemicals. Predictors of construction site accidents were mainly human factors and site conditions.

**Conclusion:** The study established the types of accidents and their predictors The types of accidents established can cause immerse human suffering and even death. Therefore, effective accident prevention policies need to be devised, adhered to and continuously reviewed.

## INTRODUCTION

In poor countries like Zambia the construction industry activities are mainly labour intensive. The industry workers are also unskilled, emigrate outside and migrate within the country. The construction work site and its environment changes daily, resulting in safety and health management in the construction sector difficult [1].

The construction industry is perceived to be a sector with an elevated level of occupational accidents and utilization of different contractors and sub-contractors on single sites. The sector is also been known to be untidy and entails tough work. The work is usually risky and bring about safety hazards which if not controlled and averted may maim, injure, or even make workers ill [2].

Chances of an accident resulting in critical effects rises if automobiles, ladders, structures or scaffolds are part of the accident [3]. A lot of companies do not administer proper and correct distinct scaffolding equipment training to its workers. A greater majority of trained personnel in scaffolding lack sufficient knowledge to detect and know when it is risky to operate a scaffold [4].

### Statement of the Problem

The construction sector is the third highest occupational related cause of morbidity in Zambia at 15%; with Lusaka province being the highest at 49.1% [5]. With such alarming levels of accidents, literature is still limited in this area, particularly on the prevalent accident types and their predictors. This study was therefore very important in that it has added to the body of knowledge and would help in progressive policy development to save lives of the construction workers.

### Objectives

To establish the types of occupational accidents and their predictors at construction sites in Lusaka city.

## METHODOLOGY

### Research Design

This study followed a qualitative research approach using an ethnographic research design because of the value of depth of data, the use of words and observations which preserved the storied lived experiences of construction workers. It explored the views, experiences, and beliefs of construction workers on the factors which contribute to the occurrence of accidents at construction sites. The study collected and appreciated the predictors that contribute to construction accidents in Lusaka city from the construction workers’ responses at a given point in time.

### Study Population

The study population comprised of construction workers who had been involved in at least one construction accident. This population was chosen because it was directly involved during the process of construction. Construction workers working on small projects such as house building and working in isolation were not part of this study.

### Sample Size

A sample size of seven participants was used because the study population was small, and the interviews generated in-depth qualitative data which required ample time to process. The more specific the characteristics of the participants in relation to the research objectives and the richer the dialogue in the interviews, the smaller the sample size can be [6].

### Sampling Strategy

Purposive and convenience sampling were used to select participants who had been directly involved in at least one construction accident before. Three construction sites were sampled conveniently by reason of proximity to the researchers. Seven participants were subsequently drawn from three different construction sites purposively. At site 1, 3 out of 21 workers were selected. At site 2, 2 out of 16 workers were selected. And at site 3, only 2 out of 30 workers were selected.

### Data Collection

Data was collected by interviewing construction workers using a semi-structured interview guide with a voice recording device. The data from the voice recorder was then transcribed. Firstly, the interview schedule was checked and tested for validity, reliability, and completeness to ensure collection of proper data. To maintain anonymity of the participants, serial numbers were used for record keeping purposes and not names. Open-ended questions in the interviews were used to get deep understanding of the problem.

### Data Analysis

Analysis of data was carried out manually by employing thematic qualitative data analysis [7]. The data was reduced before connecting it into themes and generating sub-themes to allow for the exploration of meaning and get a deeper insight of the data set in its entirety [7,8].

## FINDINGS

The findings of this research are structured based on three principal study objectives. Original narrations of participants are cited to maintain the authenticity and richness of data. Demographic details of participants are presented in table 1 below.

**Table 1:** Participant’s Demographic Details

| **Age group**<br>**(Years)** | **Pseudonym** | **Gender** | **Highest**<br>**qualification** | **Work**<br>**experience in**<br>**construction**<br>**industry(years)** |
| --- | --- | --- | --- | --- |
| **25-30** | *RP* | *Male* | *Diploma* | *8* |
| **31-35** | *MJ* | *Male* | *certificate* | *8* |
|  | *LP* | *Female* | *Diploma* | *16* |
| **36-40** | *-* | *-* | *-* | *-* |
| **41-45** | *OT* | *Male* | *Certificate* | *20* |
| **46-50** | *GM* | *Male* | *Certificate* | *15* |
|  | *CB* | *Male* | *Certificate* | *17* |
| **51-55** | *WZ* | *Male* | *Certificate* | *More than 30* |

### Summary of Study Findings

Aligned to the study objective two thematic areas emerged. These include:

Theme 1: Types of occupational accidents at construction sites in Lusaka city

Theme 2: predictors of occupational accidents at construction sites in Lusaka city

A summary of the themes and sub-themes is presented in Table 2 below:

**Table 2:**
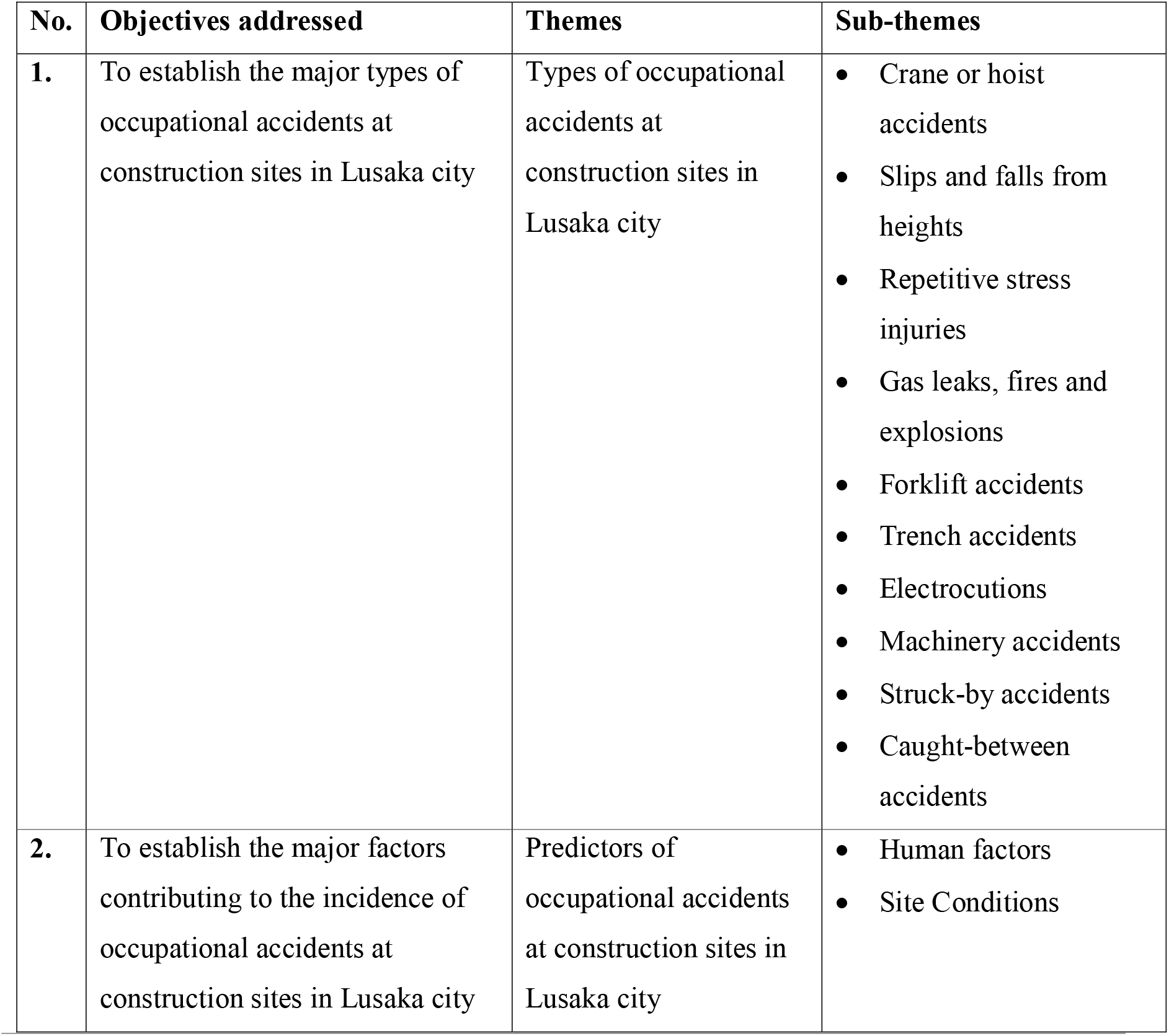
Summary of Study Findings

| No. | Objectives addressed | Themes | Sub-themes |
| --- | --- | --- | --- |
| 1. | To establish the major types of occupational accidents at construction sites in Lusaka city | Types of occupational accidents at construction sites in Lusaka city | • Crane or hoist accidents<br>• Slips and falls from heights<br>• Repetitive stress injuries<br>• Gas leaks, fires and explosions<br>• Forklift accidents<br>• Trench accidents<br>• Electrocutions<br>• Machinery accidents<br>• Struck-by accidents<br>• Caught-between accidents |
| 2. | To establish the major factors contributing to the incidence of occupational accidents at construction sites in Lusaka city | Predictors of occupational accidents at construction sites in Lusaka city | • Human factors<br>• Site Conditions |

#### Theme 1: Types of Occupational Accidents at Construction Sites in Lusaka City Crane or Hoist Accidents

Participants pointed out that crane accidents were one of the most common accidents on construction sites.

> *“As you know, cranes are mostly the biggest piece of machine in the construction environment… from my experience, a substantial number of accidents are as a result of cranes compared to any other piece of equipment at site due to malfunctions and in many accidents involving cranes or hoists, cranes collapse or buckle from too much weight. You know… sometimes construction managers ignore the fact that cranes have specifications regarding their maximum weight load*.*” (Mr. GM)*

### Slips and Falls from Heights

Respondents described three types of falls according to their experience, namely, falls on a single storey, swing falls and falls to a lower level.

> *“Fall on a single storey, mainly on footways that are not even, or which are barged in by machines, or is made of curved surfaces … worn footwear also contributes to this*.*” (Ms. LP) “…falls to a lower level from an elevated one*.*” (Mr. MJ)*
>
> *“Workers fall from a raised platform while attached to a fall arrest machine and the point of attachment to the system is usually parallel to the worker’s roof, and because the point of attachment is parallel, the lanyard that the worker is connected to makes a sharp angle… so this angle of the lanyard causes the worker to swing backwards toward the point of attachment during a free fall” (Mr. GM)*

### Repetitive Stress Injury

Accident injuries developed gradually during regular workdays caused by months and years of cumulative repetitive trauma were described by respondents.

> *“Repeated lifting, pushing wheelbarrows, pulling, vibrations from machines, or holding an awkward body position result in painful body injuries…” (Mr. CB)*

### Gas leaks, fires, and explosions

Respondents largely attributed gas leaks, fires, and explosion accidents on construction sites to negligence by workers. They described most toxic and hazardous gases as invisible to the naked eye and felt that this single property of these substances made them dangerous as they could not be detected easily using the naked eye.

> *“There is not enough time to protect oneself as explosions and fires almost always come as a surprise and. While workers that are not near could be injured by flying debris, workers nearby might be thrown from the site of the blast, … the dangers of being physically thrown and struck by objects huge, explosions can also result in excruciating burns…plus with almost every explosion comes the risk of fire. An entire structure can collapse due to extreme explosions*.*” (Mr. RP)*

Participants described some toxic and hazardous chemicals as completely unavoidable.

> *“While working in a confined room, I remember we were not wearing respirators, my colleagues and I developed terrible headaches after about four hours of continuous work. We did not know what was happening we only came to learn later that some carbon monoxide had leaked into the room after investigations were done*.*” (Mr. RP)*

### Forklift accidents

Forklift accidents were named as one of the accidents that occur frequently at construction sites.

> *“You know… a forklift as a piece of heavy machinery can be extremely dangerous under certain conditions. It is a powered industrial truck designed to move heavy materials and supplies*.*” (Mr. GM)*

### Trench accidents

Respondents stressed that trenches, which are frequently utilized at most sites of construction in Lusaka to cover plumbing and electrical wires or to get to areas underground can be a source of dangerous accidents.

> *“When not properly secured with a retaining wall and other structural reinforcements a trench may curve in… collapsing may become inevitable, and thousands of pounds of dirt may crush the victim … so when there is not enough respiratory protection or inadequate ventilation supplied to workers asphyxiation may occur because of toxic fumes and lack of oxygen*.*” (Mr. RP)*

### Electrocutions

Participants shared the view that construction workers who are exposed to electrical current at construction sites can suffer several bodily injuries such as flash burns or arc burns, electrical burns, shock and thermal contact burns.

> *“Electricity is really not friendly, direct exposure to it causes electrical accidents such as coming into contact with an underground power line while excavating, like what happened to me five years ago… inability to identify and meeting energised torn or naked cables, faulty electrical machine or gadgets. I can say at construction sites… categories of accidents involving electricity are in three groups… and I think coming in contact with overhead power lines is the most frequent type of accident. The second type comes about when workers come into contact with live cables and transformers. In the third type, while working with machinery, tools and other equipment…one can come in contact with electrical current*.*” (Mr. WZ)*

### Machinery accidents

Participants narrated how they had been involved in machinery related accidents.

> *“I remember, uh, uh, my first accident involved faults in assembly of the machine I was using to level rough surfaces. This was discovered during post-accident investigations, uh, it was discovered that the machine was not properly assembled when it was undergoing routine maintenance. The tip of my little finger was crushed when a part of the machine I was using fell off because it was loose*.*” (Mr. OT)*
>
> *“At one time while working, my trousers got caught by a front loader machine which was being operated by a colleague… It was bad for my friend who was run over and later died at the hospital, but I was lucky I survived with a few bruises*.*” (Mr. CB)*

### Struck-by accidents

Participants felt that struck-by accidents usually cause a cascade of other accidents when they occur at site and that safety gear is very helpful.

> *“Large metal pipes came rolling towards us, we were about seven in number that afternoon on the ground floor of the structure we were building we could hear the sound of them bumping into each other. Fortunately, we narrowly managed to get out of their way and that was how we escaped unhurt, though parts of the building which were hit by them got broken because the impact was so much, there were huge vibrations we almost thought the entire building was going to collapse, uh, uh, it was so scary. So, because of the impact a lot of objects from the top floors came down flying, and had it not been for the hard hats we were wearing plus safety boots we would have sustained serious injuries. Most of the materials that were being mechanically lifted swayed and fell” (Mr. OT)*

### Caught-in between accidents

Participants pointed out that if machinery is not properly guarded or is not locked down when not in use, the machinery can accidentally turn on and workers can get their clothes or body parts caught between the machinery.

> *“We were just starting our project and an excavator had just been brought on site when it accidentally turned on and we were almost caught in between its parts because we were resting just below it since there was some good shade*.*” (Mr. CB)*

### Predictors of occupational accidents at construction sites in Lusaka city

Participants described various factors they felt contributed to the occurrence of accidents at sites of construction.

### Human factors

Participants observed that the risk of fatigue-related accidents at construction sites increased with increased number of shifts done in a week.

> *“I remember the project we were working on was behind schedule so we could work from morning till evening for more than two weeks, weekends inclusive. It became physically, mentally, and psychologically very demanding on our part as workers. We rarely took enough meal breaks or rest between demanding tasks… the number of injuries amongst us increased exponentially*.*” (Mr. GM)*

Lack of training was seen as a contributing human factor to accident occurrence at construction sites.

> *“So, I think lack of training to identify signs of drowsiness, adhere to mandatory shift breaks, and education on best practices to reduce fatigue on the job greatly contribute to accidents*.*” (Mr. MJ)*

Participants strongly felt that workers who abused alcohol and other drugs were at higher risk of being involved in an accident.

> *“I strongly feel construction workers who abuse alcohol and other drugs are usually at risk of dropping materials, falling, indiscriminately abusing electrical machines, and committing all sorts of errors that can harm everyone around the construction site and themselves too*.*” (Ms. LP)*

### Site Conditions

Unstable working surfaces and restricted mobility especially in high up working places at construction sites were among factors which respondents felt contributed to the occurrence of construction accidents.

> *“Unstable working surfaces such as mud or sand uh, uh, and I think even bulk storage of materials that do not provide a firm footing such as rubble materials almost always lead to accidents, and when these unstable working conditions are off the ground it becomes very dangerous. Also, in some cases limited accessibility and movement at heights have been contributing to the occurrence of accidents, safety machine failure or misuse, plus the probability of human mistake*.*” (Mr. CB)*

Respondents described equipment that have defective designs and are sub-standard as having a high likelihood to malfunction and eventually cause accidents at construction sites.

> *“In my own opinion it has become common in recent years to be provided with sub-standard safety equipment which have defective design… they give these to us to use on construction sites for cost saving reasons. The defective ones usually malfunction when you need them most, uh, uh, imagine having failed bolts, ladders, hydraulic air lines or brake lines while working at heights*.*” (Mr. RP)*

Respondents felt that congestion and noise at construction sites could affect their response to various accident-causing hazards.

> *“When it is too crowded and noisy, it is impossible for most of us to avoid hazards such as a wrecking ball, a falling backhoe bucket, a falling ladder, a forklift blade, or turn away from flying debris, which can lead to serious personal injury*.*” (Mr. WZ)*

## DISCUSSION

### Types of Occupational Accidents at Construction Sites in Lusaka City

The types of construction accidents found to occur at sites of construction in Lusaka included hoist or crane accidents, falls and slips at heights, leaks of gas, repetitive stress injuries, explosions and fires, trench accidents, forklift accidents, electrocutions, struck-by accidents, exposure to dangerous chemicals, machinery accidents and caught-between accidents.

The study has shown that majority of the accidents which are due to machinery are caused by cranes than any other piece of equipment at site due to malfunctions such as buckle or crumble from excess weight. This outcome is consistent with preceding research in which it was found that crane accidents are perceived as one of the hazardous occurrences at sites and that cranes are indispensable as they are used to work at great heights, lift up heavy objects and in moving objects horizontally or vertically on the site [9]. The second most common type of machinery related accidents are caused by forklifts. Forklifts are extremely dangerous because they are powered industrial trucks designed to move heavy materials. Further, machine related accidents involved faults in assembly of the machines when undergoing routine maintenance. The study shows that when machinery is not properly guarded and locked down when not in use, they can accidentally turn on and badly injure workers. Serious danger is posed to construction workers when operating equipment at site with two or more moving objects and one stationary object such as a semi-trailer and a brick wall, a truck frame, and a lowering hydraulic bed.

The study shows that three common categories of fall accidents occur at sites of construction in Lusaka including single storey falls, swing falls and falls to a lower storey. Single storey falls occur mainly on footways that are not even or are encroached by equipment. This type of fall as well as falls to a lower level are attributed to worn footwear. Falls from swings happen when workers fall from a raised platform while attached to a fall arrest machine and the point of attachment to the system is usually parallel to the worker’s roof, and because the point of attachment is parallel, the lanyard that the worker is connected to makes a sharp angle, in turn this angle of the lanyard causes the worker to swing backwards toward the point of attachment during a free fall [10].

This study has shown that accidents which occur because of being struck-by objects at construction sites usually cause a cascade of other accidents when they occur, and that safety gear is very helpful to protect workers. Struck-by accidents are a regular occurrence at construction sites as evidenced by many scholars [11,12]. Trenches which are commonly used at many construction sites in Lusaka to bury electrical wires and plumbing or to gain access to underground areas are a source of dangerous accidents. Trenches cave in when not properly secured with a retaining wall and other structural reinforcement. When they collapse and result in the victim being crushed by thousands of pounds of dirt, with inadequate ventilation or insufficient respiratory protection provided to workers, asphyxiation may occur because of lack of oxygen or the presence of toxic fumes.

Further, this study shows that accident injuries developed gradually during regular workdays caused by months and in some instances years of cumulative repetitive trauma are common at construction sites in Lusaka. This type of accident is attributed to repeated lifting, pushing wheelbarrows, pulling, vibrations from machines, or holding an awkward body position resulting in painful body injuries.

The study has shown that accidents due to gas leaks, fires, and explosions on construction sites are largely because of negligence by fellow workers who do not follow safety precautions [10]. Most toxic and hazardous gases were described as invisible to the naked eye and that this single property of these substances made them dangerous as they could not be detected easily using the naked eye. The other type of accidents which occur at construction sites is caused by electrical current. This is consistent with findings in a study in Nigeria where it was found that electrocutions were among the types of accidents which occur at construction sites [13]. In certain instances, workers who are exposed to electrical current suffer several bodily injuries such as electrical burns, arc/flash burns, thermal contact burns or even shock. For example, direct exposure to electricity such as contact with an underground power line while excavating, failure to recognize energised damaged or bare wires, defective electrical equipment or power tools may result in electrical accidents. According to this study, three categories of electrical accidents at construction sites in Lusaka city are common with the most frequent type being contact with overhead power lines. The second type occurs when workers come into contact with transformers and live wires. The third most common type occurs when contact is made with electrical currents while working with tools, machinery, and other appliances.

### Predictors of occupational accidents at construction sites in Lusaka city

The study has shown that the main predictors of occupational accidents at construction sites are mainly human factors and site conditions. This finding resonates well with a study in Detroit, Michigan Kazan where it was found that the two factors responsible for accidents at construction sites are human and environmental in nature [14].

The risk of fatigue related accidents at construction sites increased with the number of hours worked per week. A study in USA also found similar results that human elements are among factors that are responsible for construction accidents [15]. The result of accidents happening due to construction projects are behind schedule and workers are made to work for extended number of hours was consistent This is coherent with findings from a study on petrochemical construction sites in the USA [16,17]. Lack of training was also seen as a contributing human factor. Most construction workers are from unskilled or not trained at all to perform construction operations and tasks [17]. It was also noted that workers who abused alcohol and other drugs were at higher risk of being involved in accidents.

Construction site conditions such as unstable working surfaces and restricted mobility especially in high up working places contribute to the occurrence of accidents. This study shows that equipment that have defective designs and are sub-standard are common and have a high likelihood of malfunctioning and eventually causing accidents at construction sites. Congestion and noise affect workers’ ability to respond to various accident-causing hazards. When it is too crowded and noisy, it becomes hard to avoid hazards such as a forklift blade, falling backhoe bucket, a wrecking ball, or turn away from flying debris or a falling ladder which can lead to serious personal injury [15]. Other studies carried out on construction accidents identified various human factors as well as site conditions as the cause [18-21]. These include: lack of enforcement of safety regulations, lack of awareness of safety regulations, poor regard for safety by people involved in construction projects; non-vibrant professionalism, engaging incompetent personnel, mechanical failure of construction equipment, Chemical impairment, emotional and physical stress.

### Study Limitations

The method of self-reported data collection is a likely constraint of this study. The conclusion would be more persuasive if more objective evidence such as site safety and health inspection records, safety and health management system audit records were reviewed. The time for the study was limited, as such I was unable to carry out repeat interviews with participants that could have helped to delve deeper into their lived experiences.

## CONCLUSION

The main objective of the study was to investigate the factors contributing to occupational accidents at construction sites in Lusaka city. Both human and site condition factors contribute to the occurrence of occupational accidents at construction sites due to gaps in the adherence to health and safety regulations. The study shows that management commitment to safety regulations is very low and this has impacted negatively on safety policy implementation. This is reflected in the various types of accidents observed at construction sites.

## Data Availability

All data produced in the present study are available upon reasonable request to the authors

## ETHICS APPROVAL AND DISSEMINATION

Ethical approval was granted by the University of Lusaka, medical Ethics Committee, this included a data extraction tool and data management plan to ensure adherence to general data Protection and health research regulations. The end study results will be published in an open access peer-reviewed journal.

## AUTHOR CONTRIBUTIONS

PS

- Work conception. Data acquisition, analysis and interpretation.
- Manuscript drafting.
- Approval of final manuscript.
- Accountable for all aspects of the work regarding its accuracy or integrity.

MZ

- Manuscript drafting.
- Critical revisions for intellectual content.
- Data interpretation
- Approval of final manuscript.
- Accountable for all aspects of the work regarding its accuracy or integrity.

RM

- Manuscript drafting.
- Critical revisions for intellectual content.
- Data interpretation
- Approval of final manuscript.
- Accountable for all aspects of the work regarding its accuracy or integrity.

## ACKNOWLEDGEMENTS

This article is a part of the master’s thesis submitted to the University of Lusaka in partial fulfillment of the requirements for the degree of Master of Public Health.

## APPENDICES

### Appendix A: Interview Guide

#### Interview schedule for construction employees, supervisors, and management personnel

Before starting each interview, rapport will be established between the interviewer and the interviewee. Thereafter, each participant will be briefed on the purpose of the study, its aim, how findings will be used and be given the opportunity to sign an informed consent form or refuse to participate. Each participant will then be given the freedom to withdraw from the study at any given time. After that, the researcher will ask for permission to record the interview for transcribing purposes.

Name of Contractor…………………………………….

Province…………………………………………………….

City…………………………………

Country……………………………

1. What nature of training do you have in the construction industry?…………………………..
2. How old are you?……………………………………………………………………………………………….
3. What is your position in your company?……………………………………………………………….
4. How long have you worked in the construction industry?……………………………………….
5. What do you think are the major contributing factors to most accidents in the construction industry?…………………………………………………………………………………………………………..
6. What nature of accidents have occurred at your construction site?…………………………..
7. How would you describe adherence to safety rules in your company?…………………….
8. How trained are the safety officers in your company?……………………………………………
9. How long are the safety trainings which safety officers and employees undergo?……..
10. How would you describe your company’s enforcement of standard operating procedures (SOPs) during construction?………………………………………………………………
11. What actions are taken when an accident happens on your construction site?…………..
12. Describe the duration of your normal and overtime working shift?…………………………
13. In your own opinion, what do you think is adequate rest required by construction workers?………………………………………………………………………………………………………….
14. What is your overtime like per week?………………………………………………………………….
15. Describe a moment(s) when you were involved or witnessed an accident at your construction site?……………………………………………………………………………………………….
16. What policies does your company have that promote construction safety?………………..
17. How current are your company safety policies?…………………………………………………….
18. What scheme does your company provide for construction workers to cover for accidents?………………………………………………………………………………………………………..
19. What do you suggest should be done to reduce the number of accidents on construction sites?…………………………………………………………………………………………………………………

### Appendix B: Participant Leaflet and Informed Consent Form

**PARTICIPANT INFORMATION LEAFLET AND CONSENT FORM TITLE OF THE RESEARCH PROJECT: MAJOR FACTORS CONTRIBUTING TO THE INCIDENCE OF OCCUPATIONAL ACCIDENTS AT CONSTRUCTION SITES IN LUSAKA CITY**

PRINCIPAL RESEARCHER: PHILLIP SIMUTENDA

CONTACT NUMBER: +260968683376

#### Introductory Part

You are being invited to take part in a research project. Please take some time to read the information presented here, which will explain the details of this project. Please ask the researcher any questions about any part of this project that you do not fully understand. It is very important that you are fully satisfied that you clearly understand what this research entails and how you could be involved. Also, your participation is **entirely voluntary**, and you are free to decline to participate. If you say no, this will not affect you negatively in any way whatsoever. You are also free to withdraw from the study at any point, even if you do agree to take part.

#### Nature and Purpose of the Study

This study will be conducted in Lusaka, Zambia. It will involve 7 participants drawn from across the city of Lusaka selected through purposive or hand-picking sampling process. The study is being undertaken to assess the major factors which contribute to the occurrence of construction accidents in Lusaka city. The study is necessary for the researcher to fulfill the requirements for the award of a degree of Masters in Public Health by the University of Lusaka. It is also being done to contribute to the growth of knowledge on construction accidents in Lusaka and to influence improvements on the current construction safety legislation.

The study is qualitative and face to face interviews will be used to collect data. The interviews will be done by myself, the principal researcher. With your consent, the interview will be recorded using a voice recorder for transcribing purposes. The findings of the study will be presented using words, but identities of participants will be protected by adopting pseudo names for all participants.

#### Selection of Participants

You have been invited to take part in this study after being selected through a purposive or hand-picking sampling procedure. This is because you have been involved in at least one construction accident on the current or previous construction sites. Your selection is also motivated by the researcher’s knowledge of your ability to contribute to the study by sharing your lived experiences as a construction worker.

#### Responsibilities of Participants

Your responsibility will be to share your lived experiences through a face-to-face interview. The interview will take place at a time and venue convenient to both parties and is likely to take between 30 and 60 minutes. Unless a need arises for a follow-up interview, you will only take part in one face to face interview.

#### Benefits of taking part in the Study

There are no personal benefits accruing to you as a participant. However, it is an opportunity for you to voice your experiences and feelings as a construction worker regarding the many construction accidents in Lusaka city. While you may not directly benefit from the research findings, future construction workers may benefit through improvements that will be influenced by the findings of this study.

#### Risks for taking part in the Study

The movement from your home to the agreed interview venue may expose you to road traffic accidents. The interview may elicit some memories and/or emotions that may cause you some emotional distress. If you require support in this regard, I will facilitate your referral to the nearest health facility for counselling.

#### Access to Research Data

The data collected will be treated as confidential and will be protected. Your identity will be protected using a pseudo name or number which you are going to propose during the interview process. In terms of access to research data, my research supervisor will have access to the research findings for academic purposes.

#### Compensation for Injuries

Participation in the study is entirely voluntary and the interviews will be conducted at your workplace to minimize on travelling distance and exposure to road traffic accidents. This should minimize the occurrence of any injury to you and as such no compensation will be provided to you. However, should an injury occur to you, you will access free medical services as provided for under the National Health Insurance Scheme of Zambia.

#### Payment for Participating in the Study

Your participation in the study is for free. You will not be paid for participating in the study.

#### Declaration by participant

By signing below, I …………………………………..…………. agree to take part in a research study entitled: *Factors Contributing to the Incidence of Occupational Accidents at Construction sites in Lusaka City*.

I declare that:

- I have read or had read to me this information and consent form and it is written in a language with which I am fluent and comfortable.
- I have had a chance to ask questions and all my questions have been adequately answered.
- I understand that taking part in this study is **voluntary** and I have not been pressurised to take part.

I may choose to leave the study at any time and will not be penalised or prejudiced in any way.

- I may be asked to leave the study before it has finished, if the researcher feels it is in my best interests, or if I do not follow the study plan, as agreed to.

Signed at (place) ………………….………..…………….. on (date) …………….……….. 2021.

Signature of participant

Signature of witness

Declaration by investigator

I (name) ……………………………………………..……… declare that:

- I explained the information in this document to …………………………………..
- I encouraged him/her to ask questions and took adequate time to answer them.
- I am satisfied that he/she adequately understands all aspects of the research, as discussed above
- I did not use an interpreter.

Signed at (place) ………………….………..…………….. on (date) …………….……….. 2021.

**Signature of investigator**

**Signature of witness**

### Appendix C: Ethics Clearance and Introductory letter

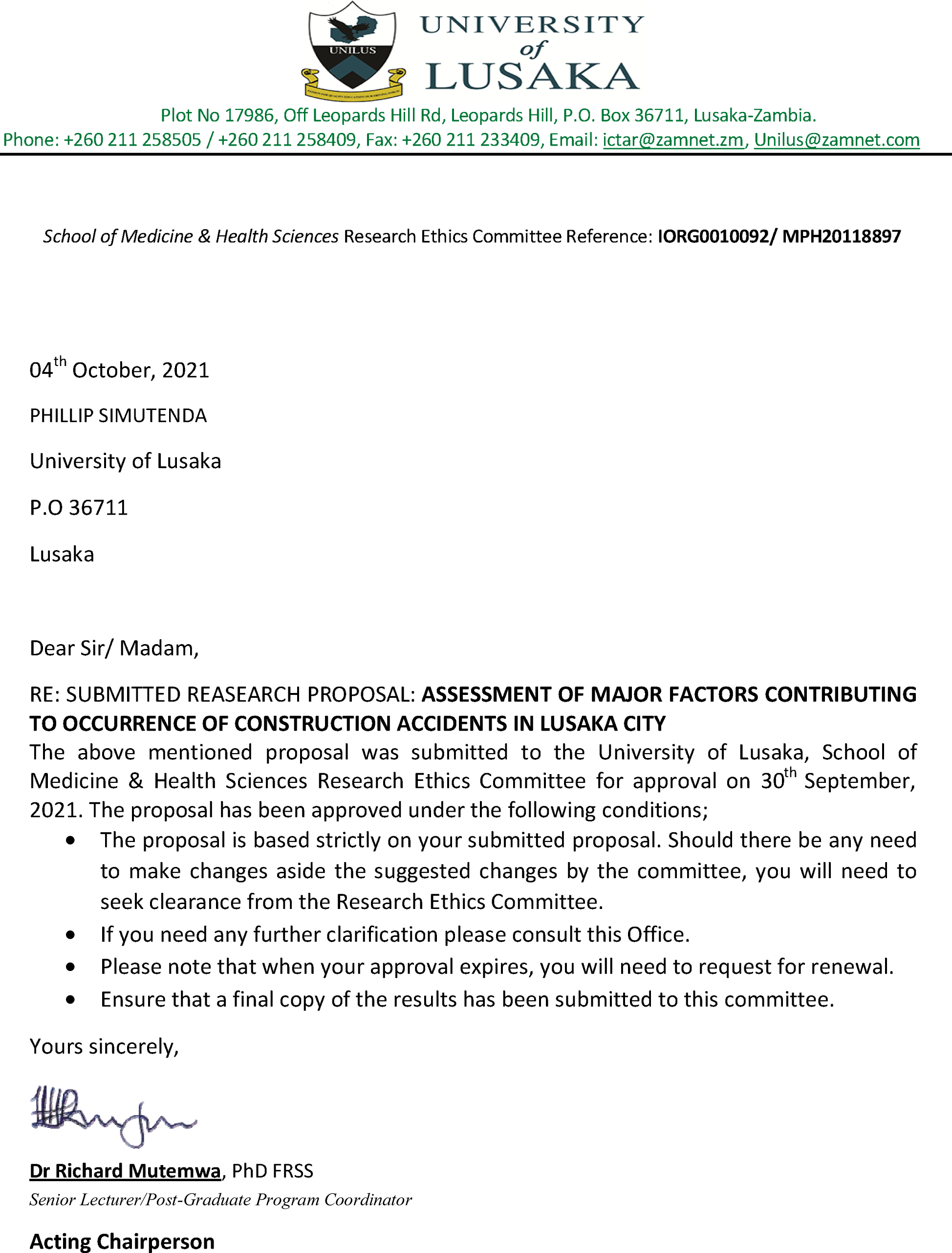

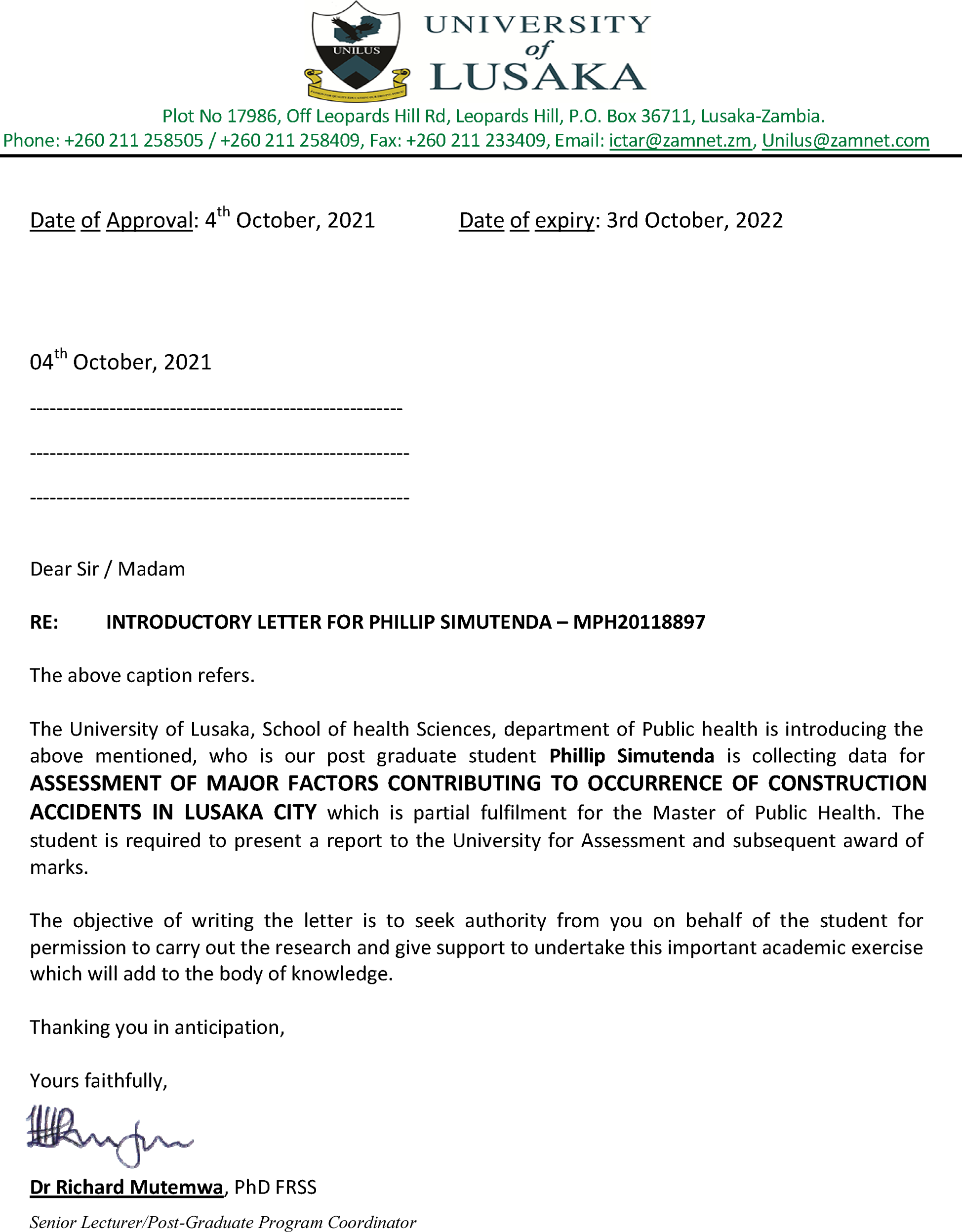

## Notes

### Competing Interest Statement

The authors have declared no competing interest.

### Funding Statement

This study did not receive any funding

### Author Declarations

University of Lusaka, medical Ethics Committee

